# The NIH Toolbox Emotion Battery and Cerebrospinal Fluid Biomarkers in Alzheimer’s Disease: Findings from the Multisite ARMADA Study

**DOI:** 10.1101/2025.05.19.25327619

**Authors:** Kexin Yu, Jennifer R. Gatchel, Emily H. Ho, Ihab Hajjar, Steven E. Arnold, Gad A. Marshall, Hiroko H. Dodge

## Abstract

**Objectives:** Investigating the relationships between socioemotional functioning and Alzheimer’s Disease (AD) pathology can contribute to screening and early detection of AD. This study explored the associations between socioemotional functioning and cerebrospinal fluid (CSF) AD biomarkers in older adults.

**Methods:** We used baseline data from the Advancing Reliable Measurement in Alzheimer’s Disease and Cognitive Aging (ARMADA) study. ARMADA is a multisite study with independent protocols for CSF assays at each site. The available sample size with comparable CSF assays had 31 participants with normal cognition (NC) and 28 with amnestic mild cognitive impairment (aMCI) or early-stage AD dementia. CSF-derived AD biomarkers included were: phosphorylated-tau 181 (p-Tau181), total tau (t-Tau), Aβ42, Aβ42/40 ratio, and p-Tau181/Aβ42 ratio. Socioemotional functioning (negative affect, psychological wellbeing, and social satisfaction) was measured with the self-reported NIH Toolbox Emotion Battery (NIHTB-EB). We ran linear regressions by cognitive subgroups (NC and aMCI/early-stage AD).

**Results:** Among participants with NC, lower social satisfaction was associated with higher p-Tau181 and t-Tau; higher t-Tau was additionally associated with more negative affect. None of the CSF AD biomarkers were associated with the NIHTB-EB outcomes among participants with aMCI or early-stage AD.

**Discussion:** These findings suggest that socioemotional functioning may be associated with tau pathology. Markers for amyloid were not related to socioemotional functioning regardless of disease stages. Future studies with larger, more diverse samples and harmonized CSF assay protocols are needed to further examine the role of early socioemotional change in early detection and prevention of dementia.

## Introduction

Socioemotional functioning refers to the capacity to experience, regulate, and express one’s emotions (Wiggins & Monk, 2013). Neuropsychiatric symptoms (NPS) describes the behavior and mood change associated with abnormal cognitive change in older adults, and can be conceptualized as negative manifestations of disrupted socioemotional functioning (Burhanullah et al., 2020). NPS are risk factors and/or prodromal conditions of Alzheimer’s Disease (AD) and have been shown to be associated with Alzheimer’s Disease (AD) pathology, such as amyloid and Tau pathology accumulation measured by both cerebrospinal fluid (CSF) biomarkers and positron emission tomography (PET) scans (d’Oleire Uquillas et al., 2018; Donovan et al., 2016; Hou et al., 2021; Krell-Roesch et al., 2018; Xu et al., 2021; Babulal et al., 2020; Gatchel et al., 2017). However, the broader role of socioemotional functioning across the trajectory of AD remains poorly understood. Examining fluctuations in socioemotional functioning—particularly those that may precede cognitive decline—can yield valuable insights into early disease processes. And such knowledge can contribute to identifying high-risk participants for AD prevention clinical trials and guide the development of innovative interventions to delay or mitigate disease progression (Donovan et al., 2016; Sperling et al., 2011).

The NIH Toolbox® for Assessment of Neurological and Behavioral Function (NIH Toolbox) Emotion Battery (NIHTB-EB) was designed to assess positive and negative emotional status as well as social wellbeing among diverse racial/ethnic and age groups and a tool for cross-group comparison (Salsman et al., 2013). To the authors’ best knowledge, no previous research has examined the associations between socioemotional function measured by NIHTB-EB and AD pathology. Identifying AD-pathology underlying socioemotional conditions is critical for early detection and prevention of Alzheimer’s Disease and Related Dementia (ADRD) (Dubois et al., 2023; Sperling et al., 2011).

Sociomeotional functioning differs aross cognitive health stages. Previous study using NIHTB-EB found older adults with mild cognitive impairment (MCI) had increased negative emotions and lower psychological wellbeing compared to those with normal cognition (NC) (Yu et al., 2022). Nonetheless, to what extent socioemotional functioning might be related AD biomarkers differ across the disease stageshas not been empirically examined. Relevant research reported the associations between NPS and AD pathology by disease stages: some empirical evidence supported such association at NC stage (Donovan et al., 2016, 2018; Gatchel et al., 2017; Krell-Roesch et al., 2018), while other reported the link between NPS and AD pathology among participants with a MCI or dementia diagnosis (Premnath et al., 2024; Tommasi et al., 2021). Building on existing work, and honing in on the borad construct of socioemotional function that may be upstream of more severe behavioral disturbances and neuropsychiatric symptoms, this study aimed to explore the associations between socioemotional constructs measured by NIHTB-EB and AD biomarkers in older adults by disease stages: with NC and those with amnestic mild cognitive impairment (aMCI) or early-stage AD diagnosis.

Investigating the associations between both positive and negative aspects of emotion, social wellbeing, and the presence of AD pathology could help develop targeted behavioral interventions for dementia prevention and improve patients’ quality of life.

## Methods

### Dataset

We used baseline data from the Advancing Reliable Measurement in Alzheimer’s Disease and Cognitive Aging (ARMADA) study. Participants in the ARMADA study were drawn from nine ongoing longitudinal Alzhemeri’s Disease Research Center (ADRC) cohorts and had been previously classified as cognitively normal, or as having cognitive impairment consistent with aMCI or early-stage dementia—both presumed to be due to Alzheimer’s disease (AD) based on clinical evaluations and, in some cases, AD biomarkers (Weintraub et al., 2022). The cognitive diagnosis was made using the 2011 NIA–Alzheimer’s Association (AA) criteria with data collected from the participants and study partners, including cognitive assessments, daily functioning assessments, neurological exams, neuroimaging, and laboratory data (McKhann et al., 2011). All sites used NACC UDS V3 for cognitive and neurological examinations (Dodge et al., 2020; Weintraub et al., 2018).

Each site had an independent protocol for CSF biomarker assessment, and there has been no valid harmonization protocol across different assay protocols (Weintraub et al., 2022). Therefore, we selected sites that had the largest sample with CSF biomarkers for each cognitive group: N=31 participants with NC from the University of Wisconsin-Madison (UW-M); N=28 with aMCI or early-stage AD from Emory University (EU) and Northwestern University (NU) with both sites using comparable CSF assay protocols (labs run by ATHENA used Luminex or Elisa methods) (Weintraub et al., 2022).

### AD biomarkers

Each site collected Cerebrospinal fluid (CSF) biomarkers data. The UW-M site used Roche Cobas 601e analyzer, while both NU and EU sites’ CSF data collection and analysis were certified by ATHENA (Weintraub et al., 2022). Five CSF-derived AD were available from the UW-M site: phosphorylated-tau 181 (p-Tau 181, pg/mL), total-tau (t-Tau, pg/mL), Amyloid-β(Aβ)42 (pg/mL), Aβ42/40 ratio, and p-Tau 181/Aβ42 ratio. The EU and NU sites have all the abovementioned biomarkers except for the Aβ42/40 ratio.

### NIH Toolbox Emotion Battery

NIHTB-EB is a socioemotional functioning assessment tool administered to participants using an iPad. For each item within subscales, participants rate on a five-point scale how often or to what extent they would agree with a statement describing socioemotional functions, with a higher score indicating more frequency and stronger agreement. Previous work identified three socioemotional domains from 17 subscales with a confirmatory factor analysis: negative affect, social satisfaction, and psychological wellbeing (*NIH Toolbox Scoring and Interpretation Guide*, n.d.). The domain scores were calculated with subscale raw scores weighted by confirmatory factor loading, and then standardized to be centered on 50 with a standard deviation (SD) of 10 (Babakhanyan et al., 2018). The negative affect domain includes subscale measurements of Anger Affect, Anger Hostility, Sadness, Fear Affect, and Perceived Stress. The social satisfaction domain includes subscales of Friendship, Loneliness, Emotional Support, Instrumental Support, and Perceived Rejection. The psychological wellbeing domain includes subscale measurements of General Satisfaction, Meaning and Purpose, and Positive Affect.

### Analysis

We conducted linear regression models with participants with NC (UW-M site) and aMCI/ early-stage AD (EU and NU) separately, with each CSF biomarker as an outcome and NIH Toolbox-EB domain scores as independent variables. To make the results across sites more comparable, we conducted a z-score transformation separately for NC and aMCI/early-stage AD groups, using their mean and SD for the CSF biomarkers. The model with EU and NU data controlled for site and disease stage (aMCI vs. early-stage AD). Additionally, all models controlled for age, sex, and education.

## Results

Table 1 summarizes the sample characteristics by NC and aMCI/early-stage AD groups. The UW-M site NC sample (N=31) had a mean age of 72.6 (*SD*=6.1), 64.5% female, 83.9% had a Bachelor’s degree, and one was African American. The EU and NU sample with aMCI or earlys-stage AD dementia (combined N=28; 20 aMCI, 8 earlys-stage AD) had a mean age of 72.2 (*SD*=5.6), 35.71% female, 82.1% had a Bachelor’s degree, and one was African American. The model results are summarized in Table 2. NIHTB-EB domain scores were only associated with CSF t-Tau and P-tau 181 in the subsample with NC. None of the CSF AD biomarkers were associated with the NIHTB-EB outcomes in the subsample with aMCI or early-stage AD (EU+NU data). Among the participants with NC, a higher CSF t-Tau was associated with higher negative affect (B=0.05, SE=0.02, *p*=.03) and lower social satisfaction (B=-0.05, SE=0.02, *p*=.01). Higher p-Tau 181 was associated with lower social satisfaction (B=-0.05, SE=0.02, *p*=.01). The Aβ-based biomarkers, including Aβ42, Aβ42/40, and p-Tau/Aβ42 ratio, were not associated with NIHTB-EB domain scores across all included cognitive stages.

**Table 1.**
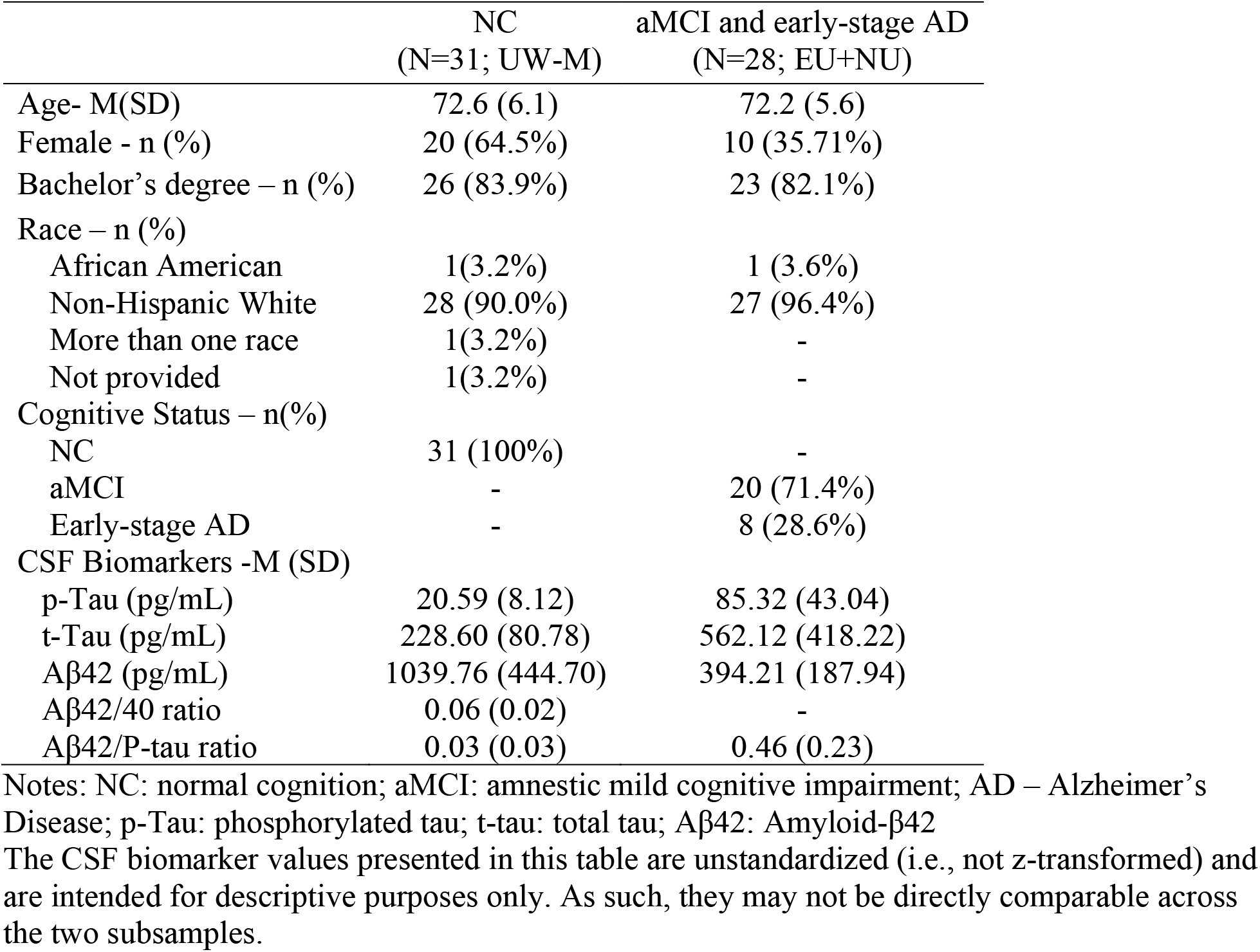
Sample Characteristics by Subsample.

|  | NC<br>(N=31; UW-M) | aMCI and early-stage AD<br>(N=28; EU+NU) |
| --- | --- | --- |
| Age- M(SD) | 72.6 (6.1) | 72.2 (5.6) |
| Female - n (%) | 20 (64.5%) | 10 (35.71%) |
| Bachelor's degree – n (%) | 26 (83.9%) | 23 (82.1%) |
| Race – n (%) |  |  |
| African American | 1(3.2%) | 1 (3.6%) |
| Non-Hispanic White | 28 (90.0%) | 27 (96.4%) |
| More than one race | 1(3.2%) | - |
| Not provided | 1(3.2%) | - |
| Cognitive Status – n(%) |  |  |
| NC | 31 (100%) | - |
| aMCI | - | 20 (71.4%) |
| Early-stage AD | - | 8 (28.6%) |
| CSF Biomarkers -M (SD) |  |  |
| p-Tau (pg/mL) | 20.59 (8.12) | 85.32 (43.04) |
| t-Tau (pg/mL) | 228.60 (80.78) | 562.12 (418.22) |
| Aβ42 (pg/mL) | 1039.76 (444.70) | 394.21 (187.94) |
| Aβ42/40 ratio | 0.06 (0.02) | - |
| Aβ42/P-tau ratio | 0.03 (0.03) | 0.46 (0.23) |

Notes: NC: normal cognition; aMCI: amnesic mild cognitive impairment; AD – Alzheimer's Disease; p-Tau: phosphorylated tau; t-tau: total tau; Aβ42: Amyloid-β42
The CSF biomarker values presented in this table are unstandardized (i.e., not z-transformed) and are intended for descriptive purposes only. As such, they may not be directly comparable across the two subsamples.

**Table 2.** Linear Regression Model Results for AD Biomarkers and NIH Toolbox Emotion Battery Domain Scores by Cognitive Status.

| T-tau | UW-M (N=31) |  |  | EU+NU (N=28) |  |  |
| --- | --- | --- | --- | --- | --- | --- |
|  | NC |  |  | aMCI+early-stage AD |  |  |
|  | B | SE | *p* | B | SE | *p* |
| Negative affect | 0.05* | 0.02 | 0.03 | 0.02 | 0.02 | 0.22 |
| Social Satisfaction | -0.05** | 0.02 | 0.01 | -0.01 | 0.02 | 0.54 |
| Psychological Wellbeing | -0.04 | 0.02 | 0.13 | -0.01 | 0.02 | 0.56 |
| P-tau |  |  |  |  |  |  |
| Negative affect | 0.04 | 0.02 | 0.08 | 0.03 | 0.02 | 0.12 |
| Social Satisfaction | -0.05** | 0.02 | 0.01 | -0.02 | 0.02 | 0.39 |
| Psychological Wellbeing | -0.03 | 0.02 | 0.24 | -0.02 | 0.02 | 0.40 |
| Aβ42 | | | | | | |
| Negative affect | 0.02 | 0.02 | 0.46 | 0.03 | 0.02 | 0.26 |
| Social Satisfaction | -0.03 | 0.02 | 0.18 | -0.02 | 0.03 | 0.40 |
| Psychological Wellbeing | -0.03 | 0.03 | 0.32 | -0.03 | 0.02 | 0.31 |
| Aβ42/40 | | | | | | |
| Negative affect | -0.02 | 0.02 | 0.35 | — | — | — |
| Social Satisfaction | 0.01 | 0.02 | 0.60 | — | — | — |
| Psychological Wellbeing | 0.00 | 0.03 | 0.87 | — | — | — |
| Aβ42/P-Tau | | | | | | |
| Negative affect | 0.02 | 0.02 | 0.45 | -0.01 | 0.02 | 0.43 |
| Social Satisfaction | -0.01 | 0.02 | 0.56 | 0.00 | 0.02 | 0.84 |
| Psychological Wellbeing | -0.01 | 0.03 | 0.83 | 0.02 | 0.02 | 0.45 |

Notes: \* $p < 0.05$ , \*\* $p < 0.01$
NC: normal cognition; aMCI: amnesic mild cognitive impairment; AD – Alzheimer's Disease; P-Tau: phosphorylated tau; T-tau: total tau; Aβ42: Amyloid-β42

## Discussion

The objective of the current study was to examine the associations between socioemotional functioning, including positive and negative emotions and social wellbeing, and AD biomarkers across cognitive stages. The findings of the current study highlighted the associations between socioemotional functioning and tau pathology in the participants with NC. We found that higher CSF t-Tau was related to lower social satisfaction and higher negative affect, and p-Tau 181 was linked to lower social satisfaction among the participants with NC. Amyloid-based biomarkers were not related to socioemotional functioning regardless of cognitive functioning and diagnosis stages.

To the authors’ knowledge, the current investigation is the first to associate NIHTB-EB with AD CSF biomarkers. Investigating the relations between socioemotional functioning and AD biomarkers has the unique advantage of upstreaming the associations between more severe behavioral disturbances and dementia pathology. In the absence of other comparable studies using the NIHTB-EB assessment, we contrasted our findings to those with NPS and AD biomarkers. Although the constructs assessed in NPS are not the same as those measured in the NIHTB-EB, key concepts of NPS, such as depressive symptoms and apathy, can be related to negative affect measured in the NIHTB-EB. Two previous studies showed increased tau PET levels were related to more depressive symptoms among older adults with NC, but no associations between Amyloid biomarkers and depressive symptoms were found (Babulal et al., 2020; Gatchel et al., 2017). On the other hand, a longitudinal investigation with 3.8 years follow-up period showed that a higher amyloid burden measured by PET imaging was related to an accelerated increase in depressive symptoms in participants with NC (Donovan et al., 2018). In participants with baseline NC, no cross-sectional associations were found between CSF biomarkers and mood, yet higher levels of baseline CSF t-Tau and CSF t-Tau/Aβ42 ratio were related to increased mood disturbance and negative emotions after a 12-month follow-up (Babulal et al., 2016). In the current study, we found associations between tau-pathology and increased negative affect in participants with NC, yet the associations between Amyloid-based biomarkers and NIHTB-EB outcomes were not statistically significant. Our findings align with prior studies in terms of the association between tau pathology and negative affect among individuals with NC (Babulal et al., 2020; Gatchel et al., 2017). However, they diverge from other reports regarding the role of amyloid burden in socioemotional functioning among older adults (Donovan et al., 2018). This discrepancy may be due to differences in sample characteristics (e.g., age, cognitive status), measurement of socioemotional constructs, or the timing of assessments relative to disease progression. Additionally, variability in the sensitivity and specificity of amyloid imaging or assay techniques across studies may contribute to inconsistent findings. Further research using longitudinal designs and larger samples with harmonized biomarker assays is needed to clarify these associations.

Related to the NIHTB-EB social satisfaction domain, a previous study did not find supportive evidence on the relationship between Amyloid-β levels and change in social engagement among cognitively normal participants (Biddle et al., 2019), but another study found individuals with higher Amyloid burden were more likely to be lonely (an indicator of low social satisfaction) in participants with NC (Donovan et al., 2016). Tau pathology accumulation measured with PET scan was also associated with greater loneliness (d’Oleire Uquillas et al., 2018). Our investigation with NIHTB-EB supports previous research findings but with different tau measures: both CSF t-Tau and p-tau were related to lower social satisfaction among the participants with NC. Amyloid-based biomarkers were not related to social satisfaction in either cognitively normal or impaired participants. It needs to be acknowledged that a few previous studies, while findings a relationship between social connectedness and cognitive decline, did not observe associations between social connection and AD pathology biomarkers, such as Tau or Amyloid-beta (Van Der Velpen et al., 2024; Wilson et al., 2007). Researchers hypothesized that a lack of social connectedness can make individuals more vulnerable to neural system change that could lead to cognitive decline but is not directly related to neuropathology (Wilson et al., 2007). The nuanced relationships between p-Tau, t-Tau, and social satisfaction need to be further explored and explained in future research.

The disease stage appears to be a critical factor for the associations between AD biomarkers and socioemotional functioning. The current study’s findings support previous research with cognitively normal older adults (Babulal et al., 2020; Gatchel et al., 2017). We did not find either tau or Aβ biomarkers to be related to NIHTB-EB domains in the sample with aMCI or early-stage AD diagnosis. Neuropathological studies have shown that Aβ accumulation proceeds the appearance of tau neurofibrillary tangles at the preclinical stage (Hampel et al., 2021). Our study findings suggest that socioemotional function can be linked with tau pathology in the AD disease spectrum before the manifestation of cognitive impairment occurs.

This study contributes to the literature by examining the associations between CSF AD biomarkers with NIHTB-EB and data from multiple research sites. The findings should be interpreted considering a few study limitations. First, the NIHTB-EB is a self-report measure. Cognitively impaired individuals might lack awareness of changes in social emotional function, which might have contributed to results. Due to a small sample size, we could not run the model separately for those with aMCI or early-stage AD. Instead, individuals with those two diagnosis were combined as a group with cognitive impairment even though there could be differences in the relationship between socioemotional functioning and biomarkers in these two disease stages. Moreover, the findings are based on cross-sectional data; hence, the directionality and time sequence of the association could not be determined. The findings need to be replicated in future studies with larger, more diverse samples and harmonized CSF assay protocols to exclude possible confounding effects.

In conclusion, for NCs, we found that higher CSF t-Tau level was related to increased negative affect and less social satisfaction, and higher CSF p-Tau level was related to lower social satisfaction. However, tau biomarkers were not associated with NIHTB-EB outcomes in the subsample with an aMCI or early-stage AD diagnosis. Aβ-based biomarkers were not related to socioemotional function across all cognitive stages. These findings suggest that aspects of socioemotional functioning—including affective states and social well-being—may serve as early indicators of AD-related pathological changes. Monitoring socioemotional changes may offer added value for early screening and detection strategies in the AD continuum.

## Data Availability

All data produced in the present study are available upon reasonable request to the authors

## Acknowledgement

This study is not preregistered. We used ChatGPT 4o to edit less than 10% of the manuscript to improve writing clarity and flow. Data, analytic methods, and study materials will be made available to other researchers upon request.

